# Interpretable biomarker programs predict treatment response in lupus nephritis: patient-level validation across four regimens

**DOI:** 10.64898/2026.09.13.26362948

**Authors:** Prashant Kumar, Shreyan Nalwad, Swapnil Keshari, Abhinav Aurange

## Abstract

A large share of late-stage clinical trial failures reflects not the underlying biology of the target but the composition of the enrolled population: trials recruit patients in whom the drug cannot work. Methods that identify likely responders *before* treatment therefore address a failure mode that better target selection alone cannot.

We applied interpretable machine learning to gene-expression data from a treatment-response cohort in lupus nephritis (GSE224705; 21,914 genes across 319 samples) covering four regimens: mycophenolate mofetil (MMF), azathioprine (AZA), hydroxychloroquine (HC) and standard of care (SOC). We independently reconstructed the expression matrix and metadata, rebuilt the treatment-specific cohorts, and derived compact multi-gene *programs* that separate responders from non-responders within each treated population.

Two results follow. First, discriminative performance is strongly graded by regimen. Compact programs of five to ten genes achieved patient-level AUROC of 0.847 (MMF) and 0.866 (AZA), but only 0.718 (HC) and 0.623 (SOC); the SOC programs performed close to chance (MCC 0.119, balanced accuracy 0.555). A regimen in which response is not transcriptionally discriminable is an actionable finding for trial design rather than a null result. Second, the programs proved considerably more stable than the differential-expression lists that generated them: reconstructed counts of significant genes differed markedly from the published analysis (222 vs. 46 for MMF; 4,455 vs. 157 for AZA; 6 vs. 24 for HC; 5 vs. 11 for SOC), yet the dominant biology and the predictive performance were preserved. Programs were also non-redundant: removing a single gene (TUBB2A) from the MMF program reduced AUROC by approximately 0.17. At the pathway level, 13 cross-treatment enrichment relationships remained significant after adjustment, indicating that response landscapes are treatment-specific yet coupled.

Patient-generalisable programs of this kind offer a concrete near-term route to enrichment-style trial design, identifying before enrolment which patients a given therapy suits. Our results also caution that the number of differentially expressed genes is a poor proxy for the strength or stability of a response signal.

## 1 Introduction

Drug development fails late and expensively. A substantial fraction of that failure is not attributable to the pharmacology of the compound but to the composition of the trial population: when a cohort mixes patients who can respond with patients who cannot, a genuine effect in the responsive subgroup is diluted below the detection threshold of the trial. This failure mode is invisible to better target selection and better chemistry alike. It is addressable only by identifying, before treatment is assigned, which patients a therapy suits.

Lupus nephritis (LN) is an instructive setting for that problem. It is a severe renal manifestation of systemic lupus erythematosus, treated with several immunosuppressive regimens whose response rates are moderate and heterogeneous, and for which no established pre-treatment molecular stratifier exists. Patients are managed on regimens including mycophenolate mofetil (MMF), azathioprine (AZA), hydroxychloroquine (HC), and combinations constituting standard of care (SOC). Because clinical response is assessed only after months of exposure, patients who will not respond accrue toxicity without benefit, precisely the scenario a pre-treatment predictor would avert.

The conventional route from transcriptomics to such a predictor runs through differential expression: identify genes that separate responders from non-responders, then use the top-ranked genes as classifier features. That route has an underexamined weakness. Differential expression output is sensitive to cohort definition, preprocessing and multiple-testing convention, and the number of genes clearing a significance threshold is routinely reported as though it were a stable property of the biology. If the feature-selection step is unstable, the predictor built on top of it inherits that instability, unless the predictive layer is empirically better conditioned than the layer beneath it.

Here we test both halves of that proposition on a public LN treatment-response cohort. We independently reconstruct the dataset, rebuild the treatment-specific cohorts, derive compact multigene programs for each regimen, and validate them at the patient level rather than reporting training performance. We then ask directly how much of the result survives when the differential-expression layer does not reproduce.

### Terminology

We use *program* throughout to denote a compact, jointly selected multi-gene feature set that is interpreted as a coherent biological module. We do not claim that the member genes are co-regulated, and we make no co-expression argument; the claim is that the genes are *jointly* informative for response, which we test directly by ablation.

## 2 Results

### 2.1 Independent reconstruction of the cohort recovers two metadata defects

We reconstructed the expression matrix and sample metadata from GSE224705 independently of the original analysis, obtaining a matrix of 21,914 genes across 319 samples and a metadata table of 319 samples by 20 fields, with complete bidirectional correspondence between matrix columns and metadata rows. The cohort comprised 299 SLE and 20 healthy samples; the SRI-4 response field was recorded as YES for 221 samples, NO for 78, and was unavailable for the 20 healthy samples. Treatment groups were AZA (35), HC (123), MMF (84) and PHC (57), with 20 samples unassigned. Published cohort composition is given in Table 1.

**Table 1.** Cohort composition as reported in the original study.

| Regimen | Responders |  | Non-responders |  |
| --- | --- | --- | --- | --- |
|  | Patients | Samples | Patients | Samples |
| MMF | 34 | 103 | 10 | 27 |
| AZA | 11 | 24 | 9 | 30 |
| HC | 56 | 133 | 14 | 40 |
| SOC | 73 | 173 | 25 | 64 |

Two metadata defects surfaced during reconstruction, both of which would propagate silently into downstream results.

First, the SOC cohort initially collapsed to zero samples. The exclusion criterion for concomitant cytotoxic therapy was being applied to a dose field stored as character strings rather than numerics, so that no comparison evaluated as intended. Parsing the numeric dose resolved 174 samples at dose 0, five at dose 150 and one missing, across 180 candidate samples from 75 unique patients. Only one patient carried a non-zero dose. Excluding that patient and the missing-dose sample yielded a final SOC cohort of 174 samples (123 HC and 51 PHC; 35 non-responders and 139 responders).

Second, a continuous clinical variable in the MMF cohort appeared to be supported by 19 rather than 20 non-responders, raising the possibility of a cohort definition discrepancy. Inspection showed the MMF cohort contained 84 samples, 20 non-responders and 64 responders throughout; the apparent reduction arose because a single *responder* had a missing protein–creatinine ratio and the analysis applied listwise deletion for that variable alone. This was variable-level missingness, not a change in cohort membership.

Neither defect is exotic. Silent type coercion and listwise deletion altering an effective denominator are common failure modes, neither can be detected from a published summary table, and either can change a reported cohort.

### 2.2 Differential expression is treatment-specific but highly sensitive to analytical convention

Applying the original limma-based differential expression framework and covariate structure to the reconstructed cohorts (Methods) yielded markedly different counts of significant genes from those published (Figure 1a). We recovered 222 significant genes for MMF (73 up, 149 down), 4,455 for AZA (1,908 up, 2,547 down), 6 for HC (1 up, 5 down) and 5 for SOC (1 up, 4 down), against published counts of 46, 157, 24 and 11 respectively. The divergence is bidirectional: our MMF and AZA counts are substantially larger, our HC and SOC counts smaller.

**Figure 1:**
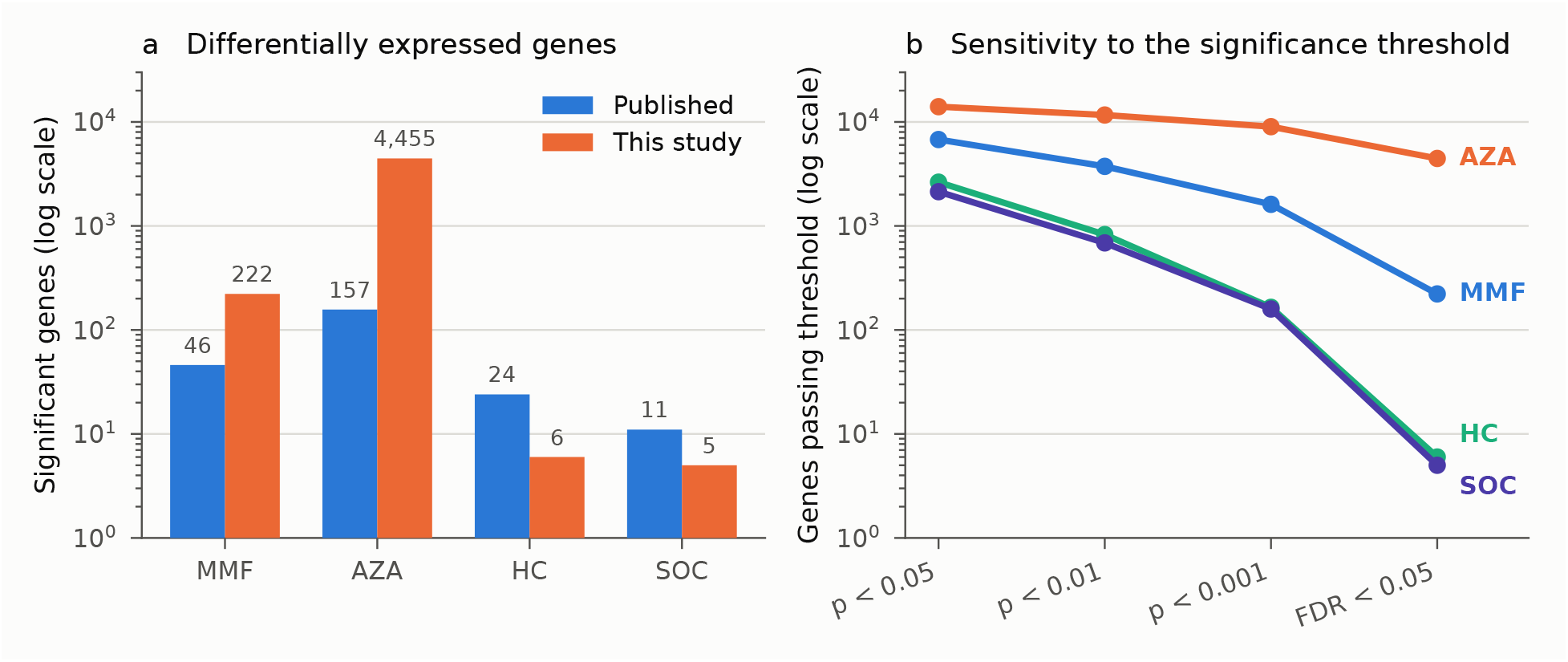
Differential expression is treatment-specific but sensitive to analytical convention. (a) Counts of significant genes per regimen, published versus this study, on a logarithmic scale. The divergence is bidirectional: our MMF and AZA counts are larger, our HC and SOC counts smaller. (b) Number of genes passing successively stricter thresholds, out of 21,914 tested. A direct Bonferroni correction returns zero significant genes for all four regimens and is therefore not plottable on a logarithmic axis.

The threshold diagnostics (Figure 1b, Table 2) show why these counts should not be treated as a stable property of the data. Across the same 21,914 tested genes, the number passing threshold changes by orders of magnitude with the convention adopted: for AZA, 14,018 genes at *p <* 0.05, 9,006 at *p <* 0.001 and 4,455 under Benjamini–Hochberg control at FDR *<* 0.05. A direct Bonferroni correction across 21,914 tests returns *zero* significant genes for all four regimens. The published analysis reports Bonferroni-significant counts of 46, 157, 24 and 11; our adjusted counts derive from Benjamini–Hochberg control. The two conventions are not interchangeable at this scale, and the gap between them exceeds the gap between any two cohort definitions we examined.

**Table 2.**
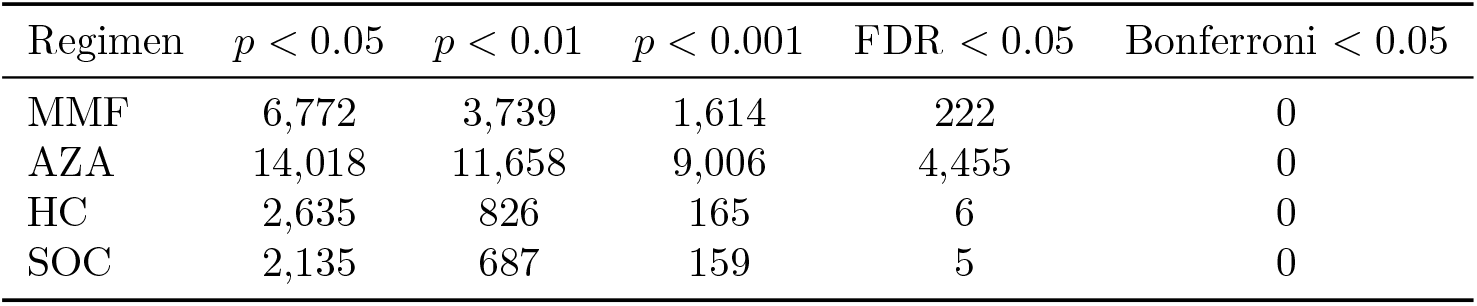
Threshold sensitivity of the differential expression result. Counts are out of 21,914 tested genes. The FDR column is the result reported throughout this manuscript.

| Regimen | $p < 0.05$ | $p < 0.01$ | $p < 0.001$ | FDR < 0.05 | Bonferroni < 0.05 |
| --- | --- | --- | --- | --- | --- |
| MMF | 6,772 | 3,739 | 1,614 | 222 | 0 |
| AZA | 14,018 | 11,658 | 9,006 | 4,455 | 0 |
| HC | 2,635 | 826 | 165 | 6 | 0 |
| SOC | 2,135 | 687 | 159 | 5 | 0 |

We note one mechanical observation without asserting a causal link: the differential-expression utility in the original codebase carries the default argument `padj=`“`bonferoni`”, a misspelling of “`bonferroni`”. We flag this because it is testable, not because we have tested it; establishing whether it accounts for any part of the divergence requires executing the original function in the original environment under both spellings.

Neither analysis is erroneous. The point is narrower: a count of differentially expressed genes, reported without the convention that produced it, carries far less information than its routine use in the literature implies.

### 2.3 Compact programs discriminate response at the patient level

We next asked whether predictive signal survives the instability documented above. For each regimen we built independent models on compact programs (ten genes for MMF and AZA, five for HC and SOC) and evaluated them at the patient level (Table 3, Figure 2).

**Table 3.** Predictive performance. Upper block: author-pipeline replication. Lower block: independent compact-program models. All metrics were computed by us; the original study reports MCC only (0.70, 0.81, 0.63, 0.56 for MMF, AZA, HC and SOC respectively). Prev. is responder prevalence in the evaluation set.

| Regimen | Model | $n$ | Prev. | AUROC | MCC | Bal. acc. | Sens. | Spec. | Acc. |
| --- | --- | --- | --- | --- | --- | --- | --- | --- | --- |
| *Author-pipeline replication* |  |  |  |  |  |  |  |  |  |
| MMF | RF | 124 | 0.847 | 0.836 | 0.626 | 0.825 | 0.934 | 0.717 | 0.897 |
| AZA | NB | 52 | 0.462 | 0.827 | 0.613 | 0.773 | 0.740 | 0.807 | 0.789 |
| HC | KNN | 182 | 0.868 | 0.818 | 0.669 | 0.807 | 0.955 | 0.660 | 0.917 |
| SOC | KNN | 271 | 0.793 | 0.634 | 0.277 | 0.606 | 0.907 | 0.305 | 0.782 |
| *Independent compact-program models* |  |  |  |  |  |  |  |  |  |
| MMF | 10 genes | 84 | n/a | 0.847 | 0.654 | 0.802 | 0.953 | 0.650 | n/a |
| AZA | 10 genes | 35 | n/a | 0.866 | 0.726 | 0.859 | 0.778 | 0.941 | n/a |
| HC | 5 genes | 123 | n/a | 0.718 | 0.410 | 0.638 | 0.981 | 0.294 | n/a |
| SOC | 5 genes | 180 | n/a | 0.623 | 0.119 | 0.555 | 0.842 | 0.268 | n/a |

**Figure 2:**
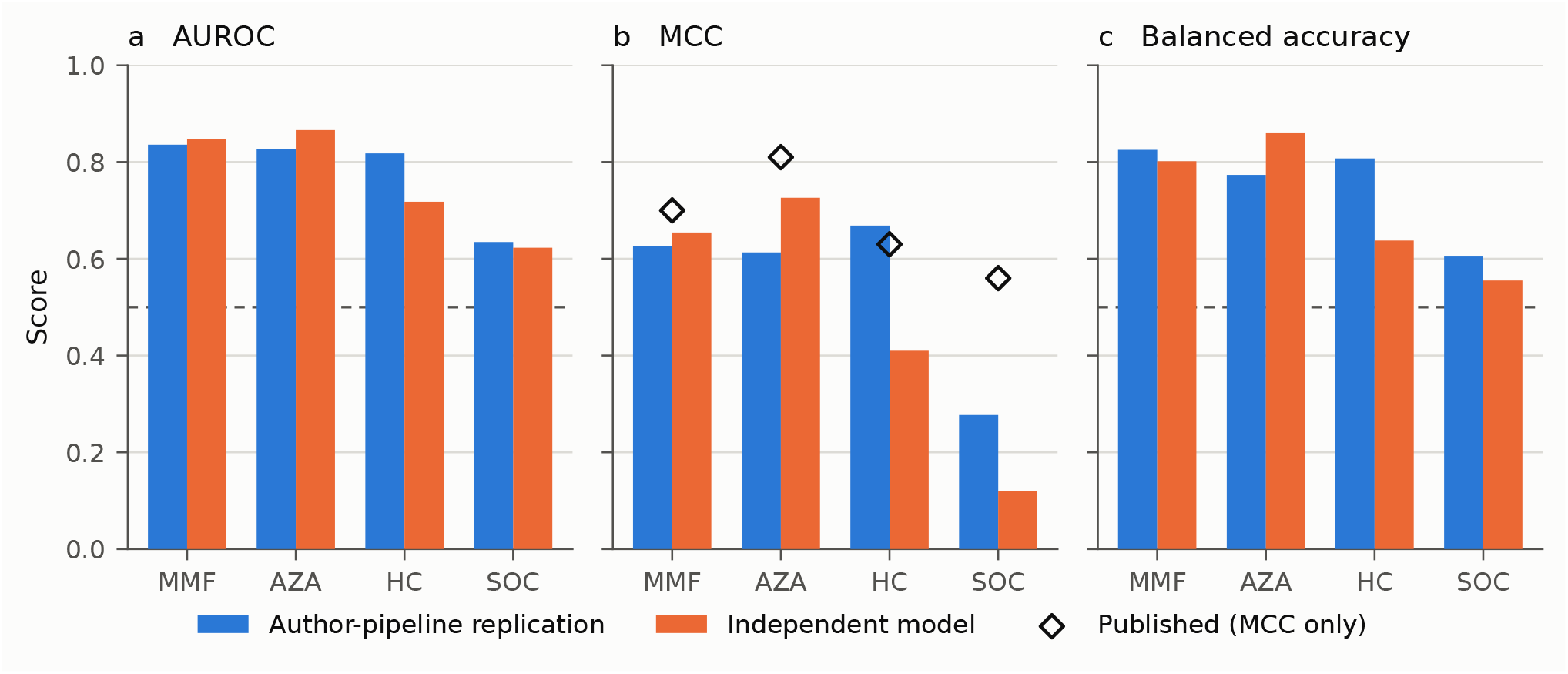
Patient-level predictive performance is graded by regimen and stable across two pipelines. Author-pipeline replication (blue) and independent compact-program models (orange) across four regimens. Dashed lines in (a) and (c) mark chance performance. Open diamonds in (b) mark the published MCC values, the only performance metric reported by the original study; all AUROC and balanced accuracy values shown are our own computation. Exact values, with class prevalence and the remaining metrics, are given in Table 3.

Performance is strongly graded by regimen. The MMF program reached AUROC 0.847 (MCC 0.654, balanced accuracy 0.802, sensitivity 0.953, specificity 0.650) across 84 samples, and the AZA program reached AUROC 0.866 (MCC 0.726, balanced accuracy 0.859, sensitivity 0.778, specificity 0.941) across 35 samples. The HC program reached AUROC 0.718 but with markedly asymmetric errors (sensitivity 0.981, specificity 0.294; MCC 0.410), and the SOC program reached AUROC 0.623 with MCC 0.119 and balanced accuracy 0.555, close to chance.

Class prevalence must be read alongside these values. Responders comprised 84.7% of the MMF evaluation set, 86.8% of HC and 79.3% of SOC, but only 46.2% of AZA (Table 3). High accuracy under strong imbalance is uninformative, which is why we report AUROC, MCC, balanced accuracy, and sensitivity and specificity separately throughout; the HC and SOC results in particular are accuracy-inflated and specificity-poor, and their headline accuracies of 0.917 and 0.782 should not be read as evidence of a usable classifier.

The programs are not merely convenient summaries of a larger signal: they are non-redundant. Removing a single constituent gene, TUBB2A, from the MMF program reduced AUROC by approximately 0.17, a degradation far larger than would be expected if the remaining genes carried equivalent, redundant information. This supports reading the feature set as a program rather than as a ranked list from which any member could be substituted. We report this single ablation as illustrative; a systematic leave-one-gene-out analysis across all four regimens is in progress and is not claimed here.

### 2.4 Predictive performance is reproducible across two independently constructed pipelines

To separate the stability of the predictive signal from the particulars of our own modelling choices, we also reconstructed the original analysis pipeline and evaluated it on the reconstructed cohorts. This replication used the algorithm families and feature-selection strategy of the original study; the original report provides MCC as its only performance metric, so all AUROC, balanced accuracy, sensitivity, specificity, precision, NPV and *F*_1_ values reported here, for both pipelines, were computed by us.

The two pipelines agree on the ordering and on the broad magnitude of the signal (Figure 2). Author-pipeline replication gave AUROC 0.836 (MMF), 0.827 (AZA), 0.818 (HC) and 0.634 (SOC), against 0.847, 0.866, 0.718 and 0.623 for the independent models. Both agree that MMF and AZA carry a substantial response signal and that SOC carries almost none. They disagree most for HC, where the independent model performs considerably worse (AUROC 0.718 vs. 0.818; MCC 0.410 vs. 0.669), consistent with HC having the weakest and least stable program of the three non-SOC regimens.

Against the published benchmark, the correspondence is partial. Published MCC values are 0.70 (MMF), 0.81 (AZA), 0.63 (HC) and 0.56 (SOC). Our replication recovered 0.626, 0.613, 0.669 and 0.277. The agreement is close for MMF and HC and poor for AZA and SOC. We therefore describe this as a pipeline replication that preserves the ranking and the qualitative conclusion, not as a numerical reproduction; the residual differences are consistent with the cohort-definition and adjustment differences documented in Sections 2.1 and 2.2. Our replication assigns SOC an MCC roughly half the published value, strengthening rather than weakening the conclusion that SOC response is not transcriptionally discriminable in this dataset.

### 2.5 The programs correspond to coherent, regimen-specific biology

The genes carrying the strongest weight in each program map onto distinct and internally consistent biological themes (Table 4). The MMF program is led by TUBB2A, FAM3B and FECH, implicating microtubule and cytoskeletal organisation, intracellular trafficking, and heme/porphyrin metabolism. The AZA program is led by DDX58, HLA-DQA1 and IFI27, implicating DNA replication and cell-cycle control, replication stress, translation and ribosome biology, and immune and antigen-processing pathways, a coherent profile for an antimetabolite acting on proliferating lymphocytes. The HC program is led by KRT72, COL6A3 and FN1, implicating extracellular matrix, integrin and fibronectin biology. The SOC program (TUBB2A, SPP1) corresponds to a limited transcriptional response with no robust pathway-level signal.

**Table 4.** Leading program genes and associated biology, with confidence grade.

| Regimen | Leading genes | Dominant biology | Confidence |
| --- | --- | --- | --- |
| MMF | TUBB2A, FAM3B, FECH | Microtubule and cytoskeletal organisation; intracellular trafficking; heme/porphyrin metabolism | Moderate |
| AZA | DDX58, HLA-DQA1, IFI27 | DNA replication, cell cycle and replication stress; translation and ribosome biology; immune and antigen processing | High |
| HC | KRT72, COL6A3, FN1 | Extracellular matrix; integrin and fibronectin biology | Low |
| SOC | TUBB2A, SPP1 | Limited transcriptional response; no robust pathway-level signal | Low |

We grade confidence in these assignments as high for AZA, moderate for MMF, and low for HC and SOC, in line with the predictive performance and the number of supporting genes in each case. The low-confidence assignments should be read as descriptive rather than mechanistic.

### 2.6 Response landscapes are treatment-specific but coupled at the pathway level

Testing each regimen’s ranked gene list against the up- and down-regulated signature sets derived from every other regimen yielded 13 relationships that remained significant after adjustment (Figure 3, Table 5). The structure is not one of simple shared response. Some pairs are positively coupled: the MMF ranking is enriched for the AZA-up signature (NES 1.451), and the AZA ranking for the MMF-down signature (NES 1.770). Others are strongly anti-correlated, most obviously SOC against HC-down (NES −1.884) and HC against MMF-up (NES −1.819). SOC participates in five of the 13 relationships, four of them negative, despite carrying the weakest individual response signal.

**Table 5.** The 13 cross-treatment enrichment relationships significant after adjustment, ordered by adjusted *p*-value.

| Ranked by regimen | Query signature | NES | Adjusted $p$ | Direction |
| --- | --- | --- | --- | --- |
| HC | AZA.down | -1.322 | $3.95 \times 10^{-8}$ | Negative |
| HC | SOC.down | -1.815 | $7.18 \times 10^{-8}$ | Negative |
| MMF | AZA.up | 1.451 | $8.42 \times 10^{-8}$ | Positive |
| SOC | HC.down | -1.884 | $8.80 \times 10^{-6}$ | Negative |
| SOC | AZA.up | -1.318 | $2.48 \times 10^{-5}$ | Negative |
| SOC | MMF.down | -1.849 | $2.48 \times 10^{-5}$ | Negative |
| AZA | MMF.down | 1.770 | $1.71 \times 10^{-4}$ | Positive |
| HC | MMF.up | -1.819 | $2.37 \times 10^{-4}$ | Negative |
| HC | SOC.up | 1.335 | $4.13 \times 10^{-3}$ | Positive |
| SOC | MMF.up | -1.555 | $8.90 \times 10^{-3}$ | Negative |
| MMF | SOC.down | -1.598 | $2.40 \times 10^{-2}$ | Negative |
| AZA | HC.down | 1.600 | $3.90 \times 10^{-2}$ | Positive |
| SOC | HC.up | 1.315 | $4.69 \times 10^{-2}$ | Positive |

**Figure 3:**
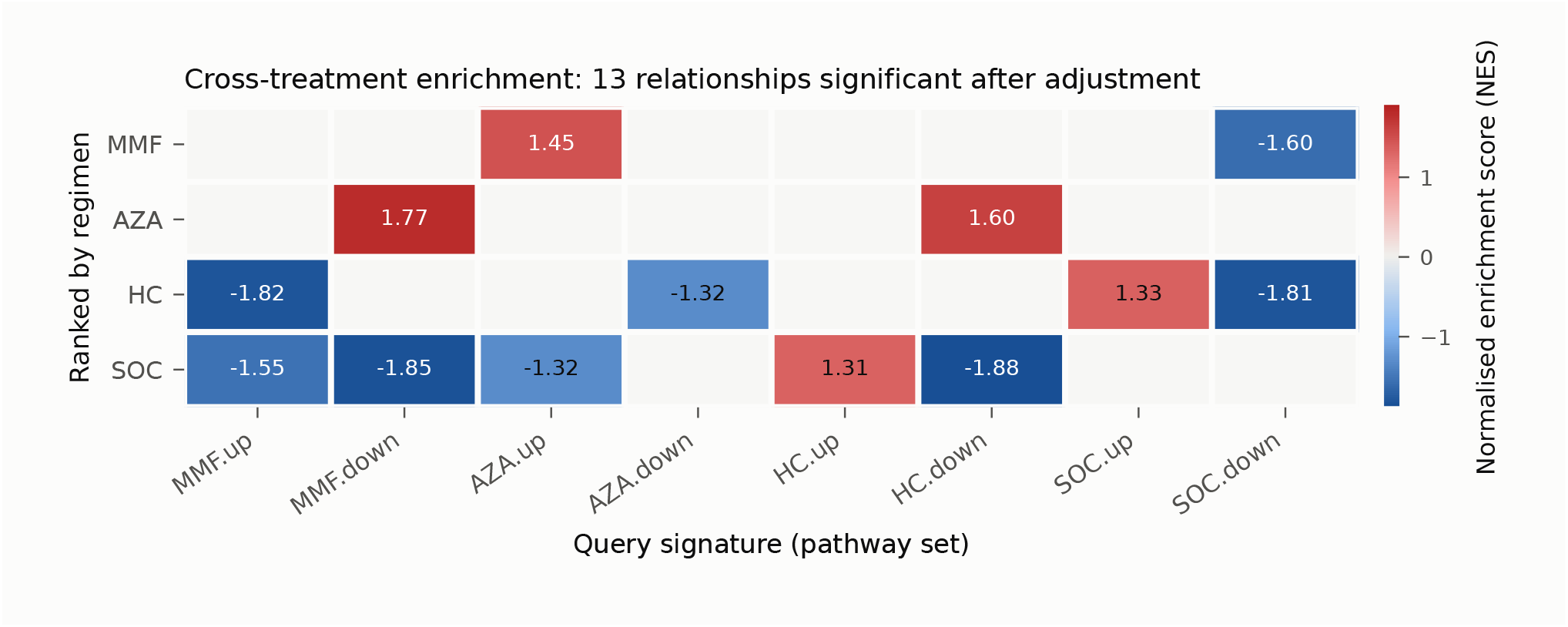
Cross-treatment enrichment relationships. Normalised enrichment scores for each regimen’s ranked gene list (rows) tested against the up- and down-regulated signature sets of every regimen (columns). Only the 13 relationships significant after adjustment are shown; blank cells were not significant. Red indicates positive enrichment, blue negative.

This is consistent with treatment-specific response programs operating on a shared underlying pathway architecture, in which different regimens engage overlapping machinery in opposing directions. It also indicates that the regimen-level signatures are not statistical artefacts of the differential expression instability documented in Section 2.2: relationships between signatures persist at pathway level even where the constituent gene lists do not reproduce in size.

## 3 Discussion

Our central result is that response predictability in lupus nephritis is graded by regimen, and that this grading is stable across two independently constructed analysis pipelines. MMF and AZA support compact programs with patient-level AUROC approaching 0.85–0.87. HC supports a weaker and less stable program. SOC supports essentially none.

The SOC result is easily mistaken for a failure of method. Under both pipelines, and across every metric, SOC response is close to undiscriminable from pre-treatment expression: AUROC 0.623 and 0.634, MCC 0.119 and 0.277, balanced accuracy 0.555 and 0.606. Two readings are available. Either response to a heterogeneous combination regimen is not encoded in the pre-treatment transcriptome at all, or SOC as defined here aggregates pharmacologically distinct treatment situations whose signals cancel. Both readings carry the same practical implication: molecular enrichment strategies should not be applied uniformly across regimens, and a regimen for which no discriminable signal exists is one where enrichment cannot help and conventional trial design remains appropriate. Knowing which regimens fall in that category before designing a trial is itself the useful output.

For the regimens where signal does exist, the translational route is enrichment. A program of five to ten genes measurable from a single pre-treatment sample, with patient-level AUROC near 0.85, is the right shape of object for a stratification assay: small enough for a targeted panel, interpretable enough to be scrutinised biologically, and validated at the patient level, which is where an enrolment decision is actually made. This is a claim about the shape of the result rather than about clinical readiness; the requirements for the latter are set out below.

The methodological finding is of independent interest. The layer at which we found least reproducibility, the count of differentially expressed genes, is the layer most often reported as a headline result. Our reconstructed counts differ from the published ones by factors ranging from 0.25 to 28, and the adjustment convention alone moves the AZA count between 4,455 and zero. Yet the biology and the predictive performance built on those lists were substantially preserved. This dissociation suggests that DEG counts function poorly as a summary statistic for the strength of a transcriptional response, and that predictive and pathway-level readouts are better conditioned targets for replication. We would encourage reporting of the adjustment convention and a threshold-sensitivity profile alongside any DEG count, as in Table 2.

Three limits on interpretation are worth stating plainly. The original analysis is not shown to be incorrect; what we show is that its gene-level output is sensitive to choices that cannot be fully recovered from the published description. Our models are not shown to outperform the published ones, because the cohorts, features and splits are not identical and the published benchmark reports only MCC. And a program validated within one retrospective cohort is not thereby shown to generalise to a prospective one.

## 4 Limitations

Several constraints bound the interpretation of this work.

### Retrospective, single-cohort design

All analyses derive from one public dataset. No external validation cohort was available, and no prospective evaluation was performed. Patient-level validation within a cohort controls for sample-level leakage but does not establish generalisation to a new population.

### Small non-responder counts

The minority class is small in every regimen: 19 non-responders in the MMF evaluation set, 28 in AZA, 24 in HC. Metric estimates on this scale carry wide uncertainty, and we report point estimates without confidence intervals because valid interval estimation requires out-of-fold probability outputs that we have not uniformly extracted across all regimens and pipelines. Comparisons between regimens should be read as ordinal.

### Class imbalance

Responder prevalence exceeds 79% in three of four regimens. Accuracy and *F*_1_ are correspondingly inflated and should not be used for cross-regimen comparison; we report them for completeness only.

### Incomplete reconciliation with the published analysis

The reconstructed cohorts are not identical to the published longitudinal cohorts, and we did not achieve exact numerical reproduction of the published DEG counts or MCC values. A line-by-line reconciliation of cohort selection, preprocessing and adjustment implementation against the original environment remains outstanding.

### Single ablation

The non-redundancy claim rests on one leave-one-out result (TUBB2A, MMF). A systematic ablation across all programs is required before non-redundancy can be asserted generally.

### Associative, not causal

The programs are correlative markers of response propensity. Nothing here establishes that the implicated genes or pathways are mechanistically involved in the response, nor that intervening on them would alter outcome.

## 5 Methods

### 5.1 Dataset and reconstruction

Expression and clinical data were obtained from Gene Expression Omnibus accession GSE224705 (BioProject PRJNA932236). The expression matrix was reconstructed to 21,914 genes by 319 samples and the sample metadata to 319 samples by 20 fields. Correspondence between expression columns and metadata rows was verified in both directions before any downstream analysis. Response was defined by the SRI-4 response field recorded in the metadata.

### 5.2 Cohort construction

Treatment-specific cohorts were rebuilt from the drug-group and dose fields. Numeric dose fields stored as character strings were explicitly parsed before any threshold-based exclusion was applied. For the SOC cohort, samples from patients with non-zero concomitant MMF or AZA dose, and samples with missing dose, were excluded, giving 174 samples from the 180 candidates. Missing values in continuous clinical variables were handled by listwise deletion within each variable; we report effective denominators per variable rather than a single cohort-level *n*, because these differ.

### 5.3 Differential expression

Differential expression used the limma framework and the covariate structure of the original study. The core routine constructs a response design matrix, removes covariate effects via `removeBatchEffect` while preserving the response contrast, fits a linear model with `lmFit`, applies empirical Bayes moderation with `eBayes`, and extracts results with `topTable`:

~~~
limma.DEG <- function(data, metadata, covars, padj) {
data <- data[, rownames(metadata)]
design.disease <- model.matrix(~Response, data = metadata)
design.covariates <- model.matrix(covars, data = metadata)
ebatch <- removeBatchEffect(data[, rownames(design.covariates)],
covariates = design.covariates,
design = design.disease[rownames(design.covariates),])
DEG <- eBayes(lmFit(ebatch,
model.matrix(~Response, data = metadata[colnames(ebatch),])))
topTable(DEG, number = nrow(ebatch), adjust.method = padj)
}
~~~

Significance was assessed by Benjamini–Hochberg control at FDR *<* 0.05. For the threshold-sensitivity analysis (Table 2) we additionally tabulated unadjusted counts at *p <* 0.05, *p <* 0.01 and *p <* 0.001, and a direct Bonferroni correction across all 21,914 tested genes.

### 5.4 Predictive modelling

Two modelling tracks were run. The *author-pipeline replication* follows the feature-selection and validation strategy of the original study, which selects the top ten differentially expressed genes by adjusted *p*-value as features and uses nested cross-validation with class-balanced outer folds, patient-level separation so that samples from one patient cannot span an outer train/test boundary, and repeated inner tuning. The *independent models* use compact programs of ten genes (MMF, AZA) or five genes (HC, SOC) with patient-level validation.

All metrics (AUROC, MCC, balanced accuracy, accuracy, sensitivity/recall, specificity, precision, NPV and *F*_1_) were computed by us for both tracks. The original study reports MCC only; no published AUROC exists for this cohort, and every AUROC in this manuscript is our own computation. Class prevalence is reported alongside every metric set (Table 3) because three of four regimens are strongly imbalanced.

Feature ablation was performed by removing a single gene from a fitted program and refitting under the same validation scheme, with the change in AUROC recorded relative to the complete program.

### 5.5 Pathway analysis

Up- and down-regulated signature sets were derived from each regimen’s differential expression result. Each regimen’s ranked gene list was then tested against the signature sets of every other regimen by gene set enrichment analysis, and normalised enrichment scores with adjusted *p*-values were retained. Relationships significant after adjustment are reported in Table 5.

## Supporting information

https://github.com/Prashantkumariitd/LNtherapy-reanalysis/blob/6f0bb51b76f382a95e86fd487c161e09ea30613f/SC-RNA_BIOLOGICAL_INTERPRETATION_SUMMARY.txt

https://github.com/Prashantkumariitd/LNtherapy-reanalysis/blob/6f0bb51b76f382a95e86fd487c161e09ea30613f/FINAL_REPRODUCIBILITY_AND_RESULTS.xlsx

https://github.com/Prashantkumariitd/LNtherapy-reanalysis/blob/6f0bb51b76f382a95e86fd487c161e09ea30613f/GSE224705_D5A_Comparative_Analysis_Report%20(1

## Data and code availability

Bulk expression and clinical data are available at GEO accession GSE224705 (https://www.ncbi.nlm.nih.gov/geo/query/acc.cgi?acc=GSE224705) and BioProject PRJNA932236. The original study’s analysis code is available at https://github.com/dtordom/LNtherapy and https://github.com/jordimartorell/pathMED. Reconstructed expression and metadata objects, per-regimen differential expression results, the complete analysis history and figure-generation code will be deposited on publication.

## Declarations

### Ethics statement

This study used publicly available, de-identified data from the Gene Expression Omnibus (GEO), accession GSE224705. No new human participants were recruited and no new biological samples were collected for this study. The study involved secondary analysis of the publicly available dataset.

### Funding

This research received no specific external funding.

### Competing interests

All authors are members of the founding team of PhaseoAI, which is listed as the institutional affiliation for this work. PhaseoAI has a commercial interest in AI-based biomedical and biomarker technologies. The authors received no funding or financial support from PhaseoAI for this study.

---

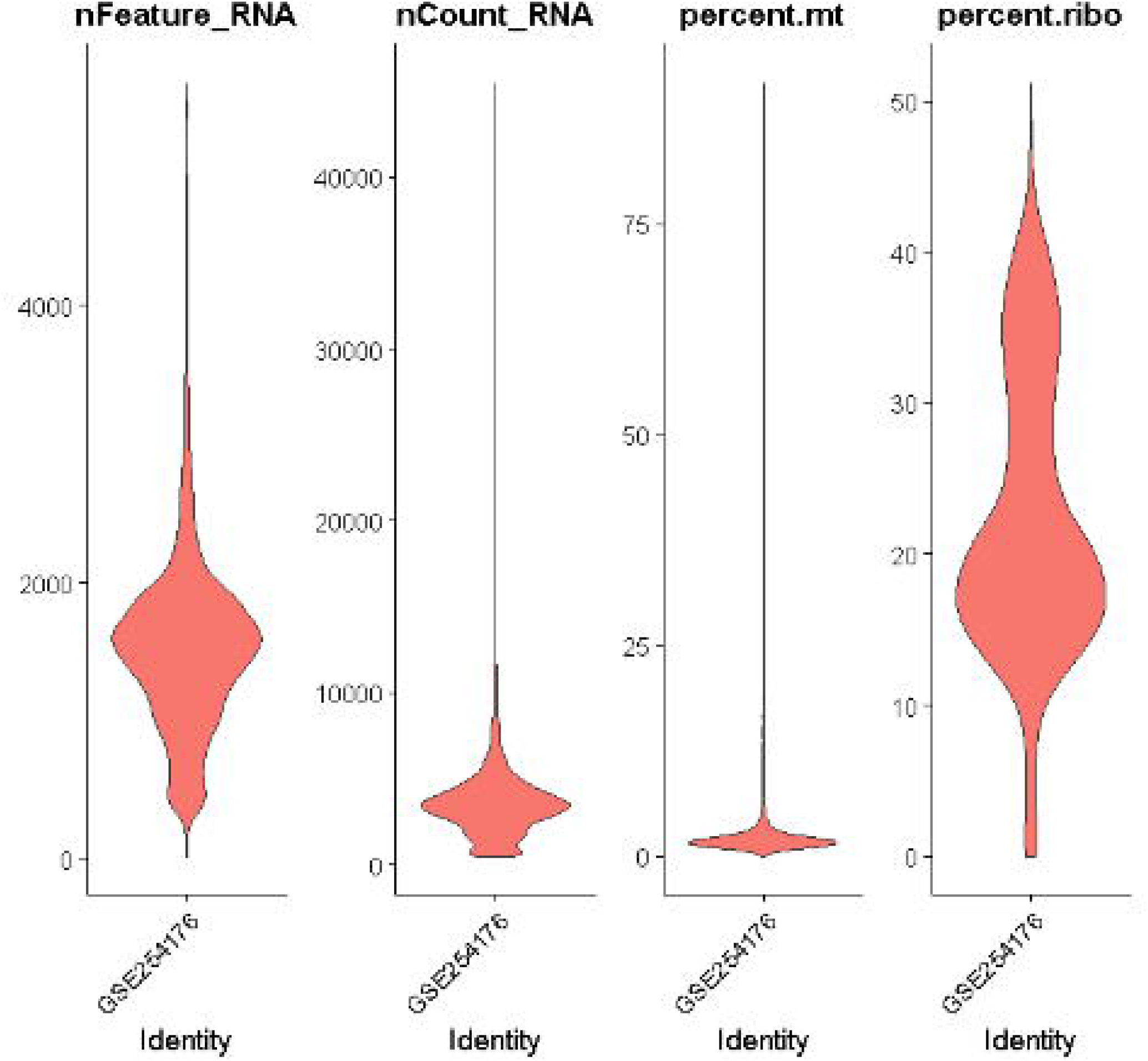

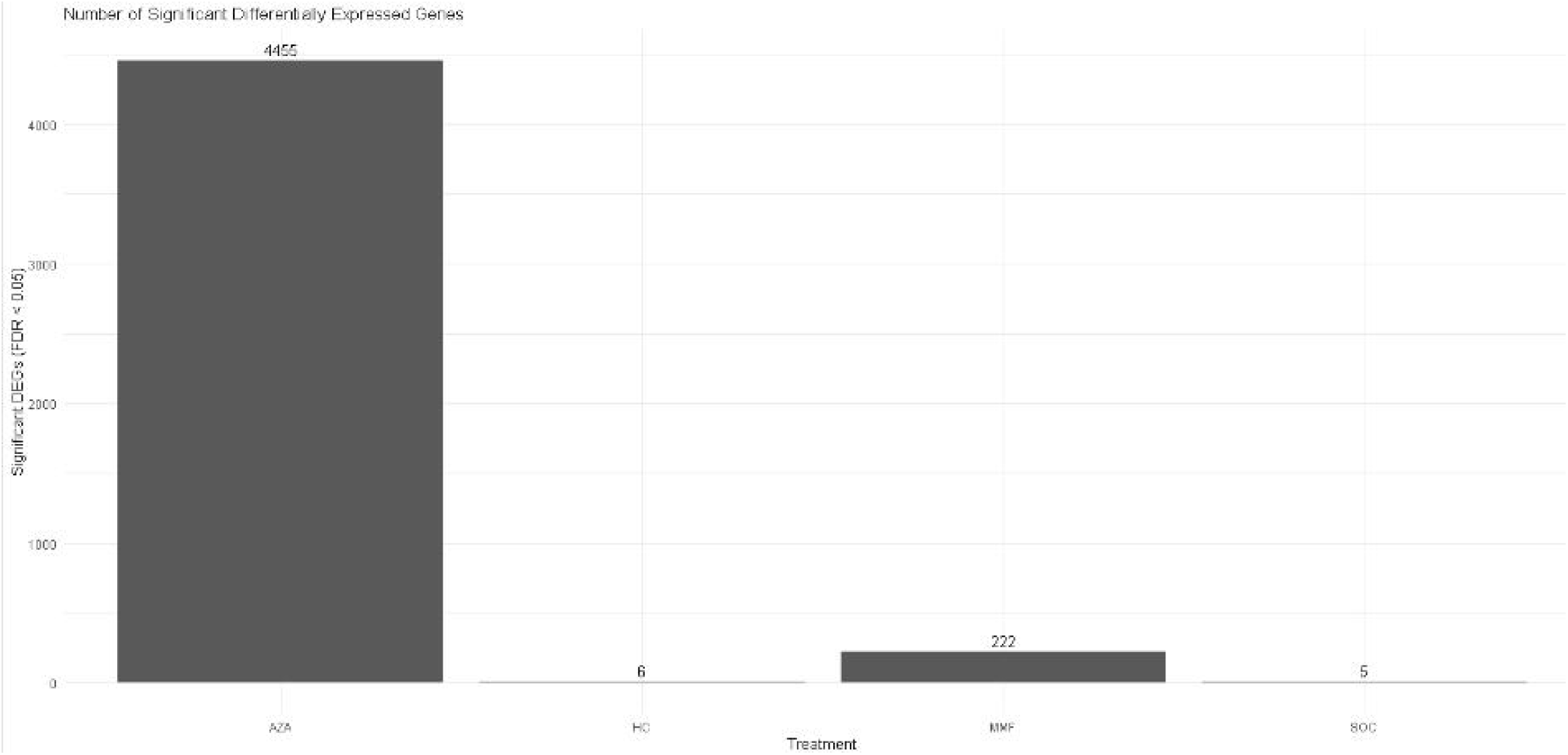

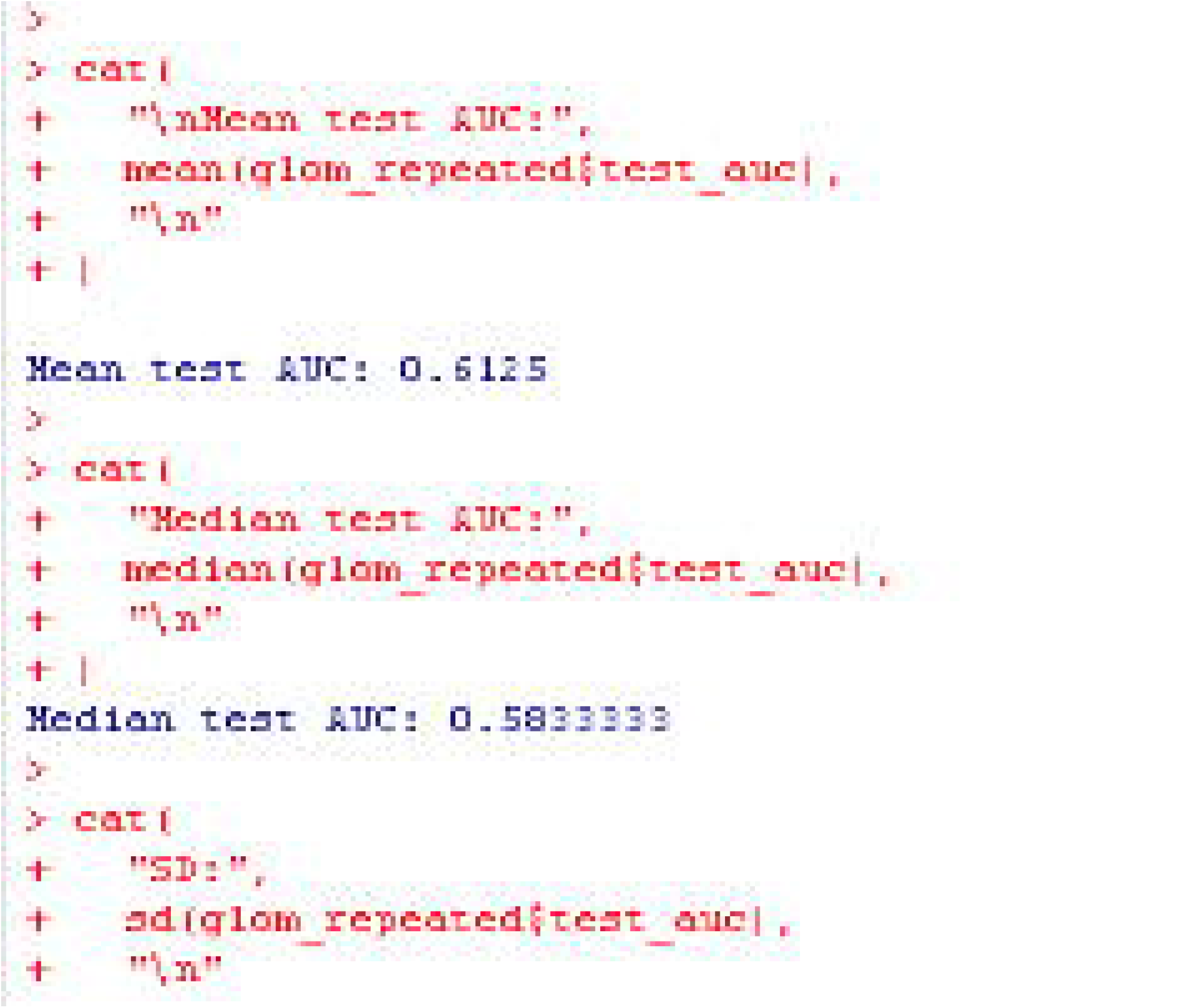

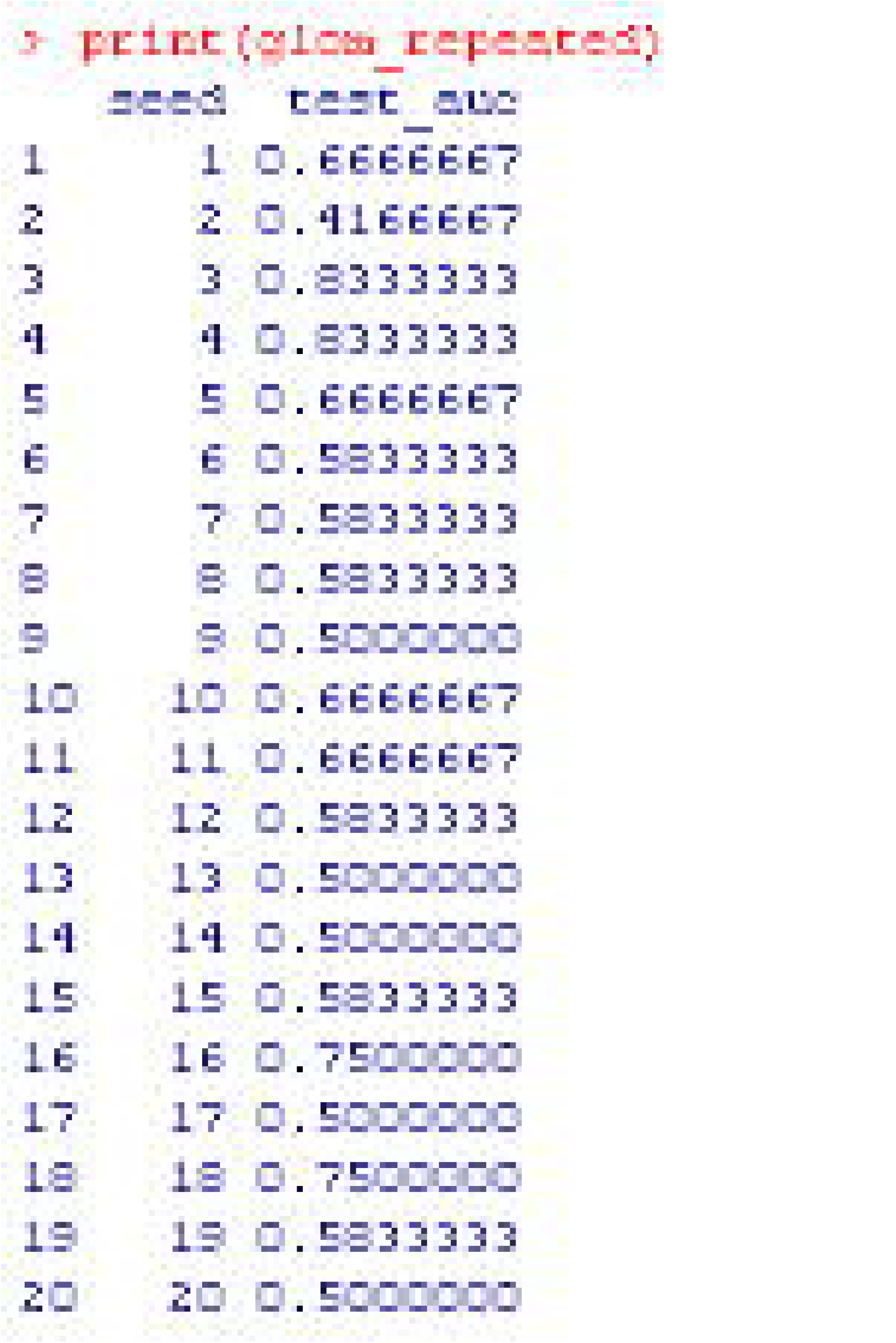

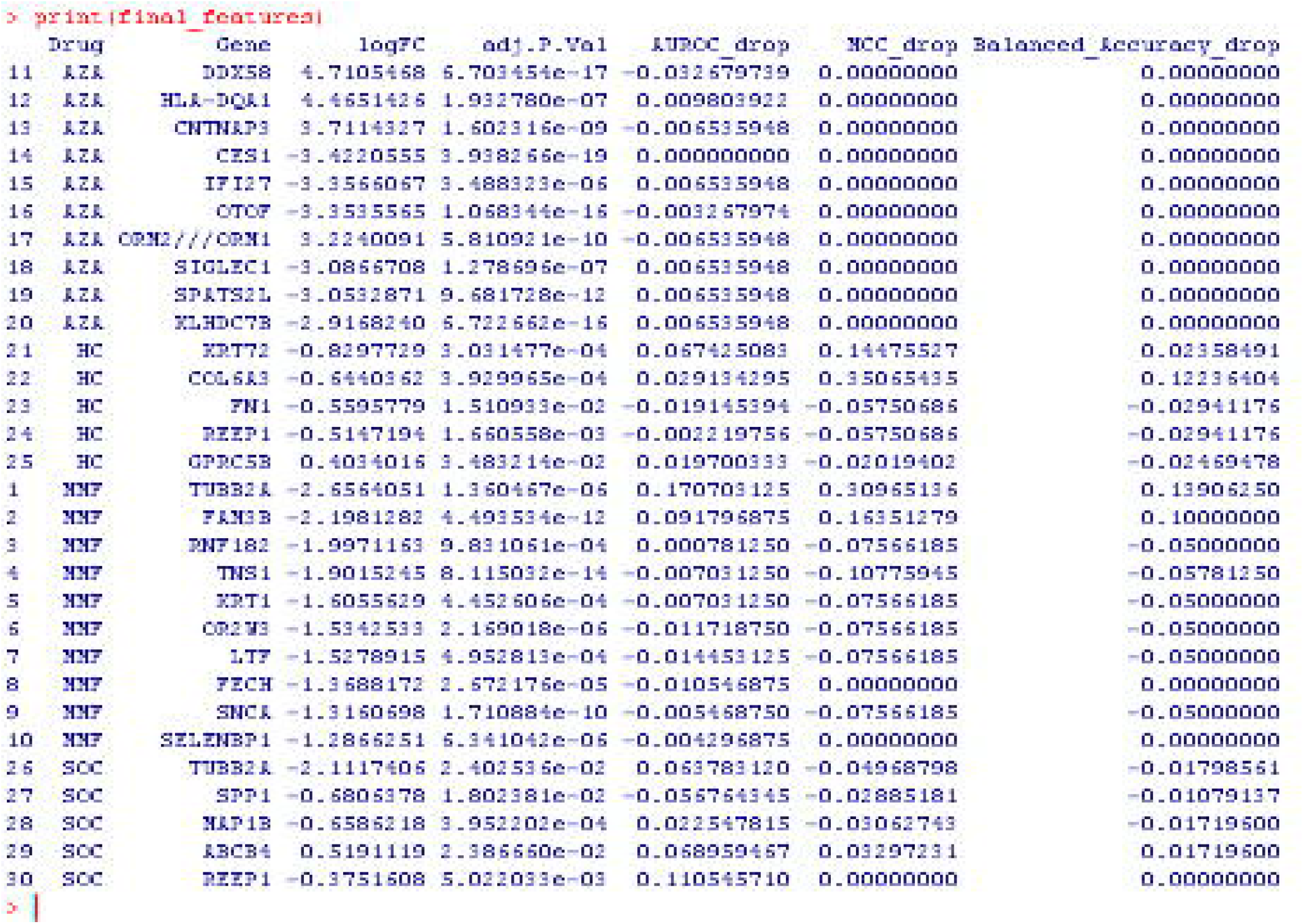

## Notes

### Competing Interest Statement

The authors have declared no competing interest.

### Author Declarations

Bulk expression and clinical data are available at GEO accession GSE224705 (https://www.ncbi.nlm.nih.gov/geo/query/acc.cgi?acc=GSE224705) and BioProject PRJNA932236. The original study analysis code is available at https://github.com/dtordom/LNtherapy and https://github.com/jordimartorell/pathMED. Reconstructed expression and metadata objects, per regimen differential expression results, the complete analysis history and figure-generation code will be deposited on publication

