## Supplementary material for "Interpretable biomarker programs predict treatment response in lupus nephritis: patient-level validation across four regimens": https://github.com/Prashantkumariitd/LNtherapy-reanalysis/blob/6f0bb51b76f382a95e86fd487c161e09ea30613f/SC-RNA_BIOLOGICAL_INTERPRETATION_SUMMARY.txt

============================================================
LUPUS NEPHRITIS scRNA-seq — PRELIMINARY BIOLOGICAL SUMMARY
============================================================
HL, HS - clinical groups
DATASET
19 biological samples: 4 HL and 15 HS.
116,701 cells and 36,601 genes.
23 annotated immune cell populations.

ANALYSIS
Cell-type-specific pseudobulk differential expression was performed.
GO enrichment and ranked-gene GSEA were then performed.
22 cell types produced significant GSEA results.

GSEA OVERVIEW
832 significant pathways in total.
431 pathways were enriched in HL.
401 pathways were enriched in HS.

MAIN HS SIGNAL
The strongest HS-associated signal occurs in antigen-presenting
and myeloid populations.

CD1C+ dendritic cells:
124 significant pathways; 106 HS-associated vs 18 HL-associated.
Strong HS programs include cytokine response, cytokine signalling,
inflammatory response, cytokine production and defense response.

CD14+/CD163+ macrophages:
78 significant pathways; 54 HS-associated vs 24 HL-associated.
Strong HS programs include inflammatory response and cytokine production.

IFN-activated dendritic cells:
61 significant pathways; 29 HS-associated vs 32 HL-associated.
HS-associated pathways include type-II interferon response,
cytokine signalling, inflammatory response and NF-kappaB signalling.

MAIN HL SIGNAL
HL shows strong pathway remodeling across lymphoid populations.
Examples include Naive B cells, Memory/atypical B cells,
Naive T/TCF7+ cells, cytotoxic gamma-delta T cells, NK cells
and plasma cells.

Cytotoxic CD8 T cells also show strong HL-associated enrichment
for chromatin-remodelling and cytoplasmic-translation programs.

BIOLOGICAL INTERPRETATION
The current results suggest that the HL and HS groups represent
distinct immune-state configurations.

HS is characterized particularly by inflammatory/cytokine/IFN-associated
programs in antigen-presenting and myeloid compartments.

HL shows a stronger lymphoid/cytotoxic and chromatin/translation-associated
program across several lymphocyte populations.

IMPORTANT CAUTION
The analysis is associative and does not by itself establish causality
or prove a renal pathogenic mechanism.

NEXT VALIDATION
Leading-edge genes should be inspected for the major inflammatory/IFN
pathways and the HL cytotoxic/chromatin programs.
The anomalous muscle/myoblast GO cluster should also be checked at
the gene level before biological interpretation.
