## Supplementary material for "Interpretable biomarker programs predict treatment response in lupus nephritis: patient-level validation across four regimens": https://github.com/Prashantkumariitd/LNtherapy-reanalysis/blob/6f0bb51b76f382a95e86fd487c161e09ea30613f/GSE224705_D5A_Comparative_Analysis_Report%20(1

**GSE224705 / D5-A
Comparative Reproduction & Analysis Report**

*Author paper vs. reconstructed analysis, methodology, results, interpretation and preprint implications*

### Executive summary

This report documents today's work on GSE224705 (D5/A) as a reproducibility-focused analysis. We first reconstructed the 21,914 × 319 expression matrix and 319 × 20 metadata, inspected the original paper and the authors' LNtherapy/pathMED code, reproduced the core author-defined limma DEG function and covariate structure, rebuilt treatment-specific cohorts, debugged the SOC metadata encoding, inspected existing ML outputs, reviewed GSEA results, and preserved the complete R history. The purpose was to establish exactly what the paper did, what we reproduced, what changed because of cohort/data reconstruction, and which extensions are justified for a future preprint.

The key result is that the core DEG framework was retained, but our reconstructed cohort produced different DEG counts from the published paper. This should be framed as a cohort/processing reproducibility question rather than as a failure or an error claim. The paper reports 46, 157, 24 and 11 Bonferroni-significant DEGs for MMF, AZA, HC and SOC, whereas our current reconstructed analysis produced 222, 4,455, 6 and 5 adjusted-significant DEGs. The exact reason for the discrepancy still requires a final line-by-line reconciliation of cohort selection, preprocessing and adjustment implementation.

| **Project item** | **Current status** |
| --- | --- |
| Dataset | GSE224705; 21,914 genes × 319 samples |
| Author method | Core limma.DEG function inspected and reused |
| DEG reconstruction | MMF 222; AZA 4,455; HC 6; SOC 5 |
| Published DEG counts | MMF 46; AZA 157; HC 24; SOC 11 |
| ML | Existing RF/NB/KNN results inspected; full paper-equivalent benchmark pending |
| GSEA | Significant cross-treatment enrichment results available |
| AUROC/AUPRC | Not yet calculated; next high-value benchmark |
| scRNA/CellChat/Hipathia | Author scripts inspected; required GSE135779-derived objects not yet recovered |
| Reproducibility | Workspace, R history and RDS outputs saved |
| R console history | 512 recorded command lines saved |

### 1. Source study and links

The source study is López-Domínguez et al., 'Immune and molecular landscape behind non-response to Mycophenolate Mofetil and Azathioprine in lupus nephritis therapy.' The paper analyzes longitudinal clinical, cellular and transcriptomic data for response/non-response to MMF, AZA, HC and SOC. GSE224705 contains expression data, drug response, doses, demographic information, visits and patient identification.

**PubMed:** https://pubmed.ncbi.nlm.nih.gov/38260685/

**Full text:** https://pmc.ncbi.nlm.nih.gov/articles/PMC10802741/

**Research Square DOI:** https://doi.org/10.21203/rs.3.rs-3783877/v1

**GEO GSE224705:** https://www.ncbi.nlm.nih.gov/geo/query/acc.cgi?acc=GSE224705

**NCBI BioProject PRJNA932236:** https://www.ncbi.nlm.nih.gov/bioproject/PRJNA932236

**Author LNtherapy code:** https://github.com/dtordom/LNtherapy

**Author pathMED code:** https://github.com/jordimartorell/pathMED

**pathMED documentation:** https://bioc.r-universe.dev/pathMED/

### 2. What the original paper reported

#### 2.1 Final cohorts

| **Treatment** | **Responder patients** | **Responder samples** | **Non-responder patients** | **Non-responder samples** |
| --- | --- | --- | --- | --- |
| MMF | 34 | 103 | 10 | 27 |
| AZA | 11 | 24 | 9 | 30 |
| HC | 56 | 133 | 14 | 40 |
| SOC | 73 | 173 | 25 | 64 |

The paper states that patients/samples receiving other immunosuppressants in conjunction with MMF or AZA were discarded, and that the final cohorts were defined after longitudinal response follow-up. SOC comprised HC and HC + glucocorticoid treatment.

#### 2.2 Published molecular and ML results

| **Treatment** | **Published DEGs (Bonferroni)** | **Published MCC** |
| --- | --- | --- |
| MMF | 46 | 0.70 |
| AZA | 157 | 0.81 |
| HC | 24 | 0.63 |
| SOC | 11 | 0.56 |

The authors used the top 10 DEGs based on adjusted p-value as ML features. Their nested validation used five class-balanced outer folds, patient-level separation so samples from one patient could not cross outer train/test sets, inner 10-fold tuning repeated five times, and 11 classification algorithms. MCC was the primary algorithm-prioritization metric.

### 3. What we reconstructed today

#### 3.1 Data and metadata

| **Object** | **Observed result** |
| --- | --- |
| Expression matrix | 21,914 × 319 |
| Metadata | 319 × 20 |
| drug_group | AZA 35; HC 123; MMF 84; NA 20; PHC 57 |
| diagnosis | Healthy 20; SLE 299 |
| sri4_response | NO 78; YES 221; NA 20 |
| Expression/metadata matching | TRUE in both directions |

```
dim(data)
# 21914 319
dim(metadata)
# 319 20
all(colnames(data) %in% rownames(metadata))
# TRUE
all(rownames(metadata) %in% colnames(data))
# TRUE
```

#### 3.2 Author code was actually inspected

We did not merely read the paper's Methods. We inspected the author repository and utility code. The central limma.DEG function uses model.matrix(~Response), removes covariate effects with removeBatchEffect, fits limma lmFit/eBayes, and returns topTable results with the requested multiple-testing method. Our DEG reproduction used this author-defined function rather than inventing a new statistical test.

```
limma.DEG <- function(data, metadata, covars, padj="bonferoni") {
    data <- data[,rownames(metadata)]
    design.disease <- model.matrix(~Response,data=metadata)
    design.covariates <- model.matrix(covars,data=metadata)
    ebatch <- removeBatchEffect(
        data[,rownames(design.covariates)],
        covariates=design.covariates,
        design=design.disease[rownames(design.covariates),]
    )
    DEG <- eBayes(lmFit(
        ebatch,
        model.matrix(~Response,data=metadata[colnames(ebatch),])
    ))
    DEG <- topTable(DEG,number=nrow(ebatch),adjust.method=padj)
    return(DEG)
}
```

### 4. DEG reproduction and paper comparison

| **Treatment** | **Paper DEGs** | **Our reproduced DEGs** | **Our UP** | **Our DOWN** |
| --- | --- | --- | --- | --- |
| MMF | 46 | 222 | 73 | 149 |
| AZA | 157 | 4,455 | 1,908 | 2,547 |
| HC | 24 | 6 | 1 | 5 |
| SOC | 11 | 5 | 1 | 4 |

These numbers are not being presented as a claim that the paper's statistical method is wrong. The important methodological point is that the same core author limma framework was used, but the reconstructed treatment-specific cohorts are not the same as the paper's final longitudinal cohorts. The paper's final sample counts and patient-selection criteria differ from the simple drug-group structure in the reconstructed 319-sample matrix. Consequently, the number of significant genes can change substantially.

#### 4.1 Significance diagnostics

| **Treatment** | **Genes** | **P<0.05** | **P<0.01** | **P<0.001** | **FDR<0.05** | **Simple Bonferroni<0.05** |
| --- | --- | --- | --- | --- | --- | --- |
| MMF | 21,914 | 6,772 | 3,739 | 1,614 | 222 | 0 |
| AZA | 21,914 | 14,018 | 11,658 | 9,006 | 4,455 | 0 |
| HC | 21,914 | 2,635 | 826 | 165 | 6 | 0 |
| SOC | 21,914 | 2,135 | 687 | 159 | 5 | 0 |

The diagnostic counts show why the adjustment convention must be frozen before a final preprint claim. The working DEG objects contain adjusted p-values yielding 222/4455/6/5 significant genes, while a separate direct Bonferroni calculation across 21,914 tests produced zero. This is a methodological reconciliation point that should be explicitly checked against the exact author environment/code path before calling the numbers a literal replication.

### 5. Clinical analysis: the 19 vs 20 issue

The MMF cohort contained 20 non-responders and 64 responders in the reconstructed clinical analysis. The apparent n=19 occurred for a particular continuous clinical variable because one responder had a missing value and the analysis used na.omit(). Thus, this was not a loss of an MMF non-responder and not a change in the cohort definition.

| **Item** | **Result** |
| --- | --- |
| MMF samples | 84 |
| MMF non-responders | 20 |
| MMF responders | 64 |
| Variable showing n=19 | protein_creatinine_ratio |
| Usable responders for that variable | 19 |
| Reason | One responder had missing value; na.omit() removed it for that variable |

resp <- na.omit(var.x[clin.tmp.x$Response=="YES"])
noresp <- na.omit(var.x[clin.tmp.x$Response!="YES"])

### 6. SOC debugging

SOC initially collapsed to zero samples because the cytotoxic-drug exclusion was applied to a dose field that was stored as character strings. Inspection showed values such as 'mmf_or_aza_dose: 0' and 'mmf_or_aza_dose: 150'. After parsing the numeric dose, 174 samples had dose 0, five had dose 150, and one was NA. Only one unique patient had a non-zero dose. After excluding that patient and the NA-dose sample, the final SOC cohort contained 174 samples.

| **SOC reconstruction checkpoint** | **Result** |
| --- | --- |
| Initial SOC candidates | 180 |
| HC / PHC | 123 / 57 |
| Unique patients | 75 |
| Parsed dose 0 / 150 / NA | 174 / 5 / 1 |
| Patients with non-zero dose | 1 |
| Final SOC samples | 174 |
| Final HC / PHC | 123 / 51 |
| Final response NO / YES | 35 / 139 |
| SOC DEGs | 5 (1 up, 4 down) |

### 7. GSEA results

The reconstructed GSEA analysis produced 13 significant treatment/pathway comparisons after adjustment. These results support the concept that the molecular response landscape is treatment-specific and that signatures from different treatments can show positive or negative enrichment relationships.

| **Drug** | **Pathway set** | **NES** | **Adjusted p** | **Direction** |
| --- | --- | --- | --- | --- |
| HC | AZA.down | -1.322 | 3.95e-08 | Negative |
| HC | SOC.down | -1.815 | 7.18e-08 | Negative |
| MMF | AZA.up | 1.451 | 8.42e-08 | Positive |
| SOC | HC.down | -1.884 | 8.80e-06 | Negative |
| SOC | AZA.up | -1.318 | 2.48e-05 | Negative |
| SOC | MMF.down | -1.849 | 2.48e-05 | Negative |
| AZA | MMF.down | 1.770 | 1.71e-04 | Positive |
| HC | MMF.up | -1.819 | 2.37e-04 | Negative |
| HC | SOC.up | 1.335 | 4.13e-03 | Positive |
| SOC | MMF.up | -1.555 | 8.90e-03 | Negative |
| MMF | SOC.down | -1.598 | 2.40e-02 | Negative |
| AZA | HC.down | 1.600 | 3.90e-02 | Positive |
| SOC | HC.up | 1.315 | 4.69e-02 | Positive |

### 8. Machine learning: what is comparable and what is not

The existing GSE224705 ML objects contain RF, Naive Bayes and KNN results. These are useful, but they are not yet a complete reproduction of the paper's 11-algorithm benchmark. The paper used top-10 DEGs and nested patient-aware cross-validation; our current stored objects need to be aligned explicitly to those features, cohort rules and split logic before a direct superiority claim is made.

| **Treatment** | **Model** | **MCC** | **Bal. accuracy** | **Accuracy** | **Recall** | **Specificity** | **Precision** | **F-score** |
| --- | --- | --- | --- | --- | --- | --- | --- | --- |
| MMF | RF | 0.6263 | 0.8252 | 0.8974 | 0.9337 | 0.7167 | 0.9443 | 0.9384 |
| MMF | NB | 0.4741 | 0.7623 | 0.7881 | 0.7979 | 0.7267 | 0.9396 | 0.8582 |
| MMF | KNN | 0.3165 | 0.6921 | 0.7494 | 0.7741 | 0.6100 | 0.9110 | 0.8347 |
| AZA | RF | 0.5463 | 0.7717 | 0.7709 | 0.7900 | 0.7533 | 0.7367 | 0.7576 |
| AZA | NB | 0.6130 | 0.7733 | 0.7891 | 0.7400 | 0.8067 | 0.8611 | 0.7595 |
| AZA | KNN | 0.5016 | 0.7467 | 0.7545 | 0.6400 | 0.8533 | 0.7029 | 0.8056 |
| HC | RF | 0.6629 | 0.7808 | 0.9277 | 0.9816 | 0.5800 | 0.9388 | 0.9593 |
| HC | NB | 0.5546 | 0.7334 | 0.9041 | 0.9668 | 0.5000 | 0.9255 | 0.9455 |
| HC | KNN | 0.6686 | 0.8074 | 0.9166 | 0.9548 | 0.6600 | 0.9502 | 0.9511 |
| SOC | RF | 0.1891 | 0.5853 | 0.7593 | 0.8827 | 0.2880 | 0.8257 | 0.8531 |
| SOC | NB | 0.1516 | 0.5653 | 0.7480 | 0.8780 | 0.2526 | 0.8169 | 0.8461 |
| SOC | KNN | 0.2771 | 0.6062 | 0.7821 | 0.9073 | 0.3052 | 0.8334 | 0.8675 |

| **Treatment** | **Paper MCC** | **Best current MCC** | **Current best** |
| --- | --- | --- | --- |
| MMF | 0.70 | 0.6263 | RF |
| AZA | 0.81 | 0.6130 | NB |
| HC | 0.63 | 0.6686 | KNN |
| SOC | 0.56 | 0.2771 | KNN |

The numerical MCC comparison is informative but not yet an apples-to-apples benchmark. The HC value being numerically higher than the published value, for example, does not establish superiority because the cohorts/features/splits are not yet identical.

### 9. AUROC/AUPRC benchmark — next high-value step

AUROC would be a strong additional benchmark because it measures ranking discrimination across thresholds. AUPRC is also valuable because response/non-response class balance can make accuracy misleading. However, AUROC must be calculated from valid out-of-fold probability predictions, not from training predictions or hard class labels. The existing MMF object contains subsample.preds and should be inspected before calculating it.

```
MMF_final <- readRDS(
  "GSE224705_MMF_ML_test_pathMED_current.rds"
)
str(MMF_final$subsample.preds)
colnames(MMF_final$subsample.preds)
head(MMF_final$subsample.preds)
```

If the prediction object contains out-of-fold probabilities, we can calculate AUROC/AUPRC and confidence intervals. If it contains only class labels, a valid ROC curve cannot be reconstructed from those labels alone.

### 10. Single-cell, CellChat and Hipathia relationship

| **Author script** | **Role** | **Today's status** |
| --- | --- | --- |
| 05_scRNASeq_mainCells.R | Main scRNA clustering/cell-type analysis | Inspected; required local GSE135779 objects not found |
| 06_scSubClustering.R | Subclustering of major cell types | Inspected |
| 07_CellCellComm.R | CellChat communication analysis | Inspected |
| 08_ApplyHipathia.R | Pathway/circuit response under target inhibition | Inspected |

The author pipeline connects bulk DEGs to single-cell cell populations, then to CellChat communication pathways and Hipathia theoretical target inhibition. Today's work did not falsely treat this downstream component as completed: the required GSE135779-derived objects were not found locally. This remains a clear next stage.

### 11. What is genuinely different from the paper?

| **Dimension** | **Paper** | **Today's work** |
| --- | --- | --- |
| Cohort | Final longitudinal, treatment-history filtered cohorts | Reconstructed 319-sample matrix with treatment-specific filtering |
| DEG method | Author limma framework | Same core author limma function and covariate structure |
| DEG counts | 46/157/24/11 | 222/4455/6/5 |
| ML features | Top 10 DEGs | Existing ML objects inspected; full top-10/patient-aware reproduction pending |
| ML algorithms | 11 | 3 currently available in inspected objects |
| Primary ML metric | MCC | MCC plus balanced accuracy, accuracy, recall, specificity, NPV, precision, F-score |
| AUROC/AUPRC | Not established as a published benchmark today | Planned |
| Mechanistic extension | scRNA + CellChat + Hipathia | Author scripts inspected; independent rerun pending |

### 12. Conclusions

- The data and metadata reconstruction is stable and saved.
- The author DEG methodology was directly inspected and used for the core reconstruction.
- The reconstructed results are reproducible and show strong treatment-specific differences in the number of significant genes.
- The difference between published and reconstructed DEG counts is a scientific reproducibility question involving cohort/sample selection, preprocessing and adjustment implementation; it should not be framed as a failure.
- The MMF n=19 issue was resolved as variable-level missingness, not a reduction from 20 non-responders to 19.
- The SOC cohort issue was resolved as a metadata encoding/parsing problem.
- The existing ML results contain useful signal, especially MMF RF, but are not yet a direct paper-equivalent benchmark.
- GSEA provides a coherent additional layer of treatment-specific molecular relationships.

### 13. What we should not claim yet

- exact numerical replication of the paper's DEG counts.
- that our current ML models outperform the paper.
- AUROC superiority before extracting valid out-of-fold probabilities.
- GSE135779 single-cell validation is completed.
- clinical utility or external validation from this retrospective public dataset.

### 14. Preprint foundation and remaining work

| **Component** | **Readiness** | **Next requirement** |
| --- | --- | --- |
| Dataset/provenance | High | Freeze versions and file checksums |
| Author-method reproduction | High | Add clean script and environment record |
| DEG comparison | Moderate-high | Resolve cohort/processing/adjustment discrepancy |
| GSEA | Moderate-high | Finalize pathways and figures |
| ML | Moderate | Exact feature/split alignment; AUROC/AUPRC + CIs |
| scRNA | Pending | Recover/regenerate GSE135779 objects |
| CellChat/Hipathia | Pending | Run from validated scRNA inputs |
| External validation | Not done | Independent cohort or prospective dataset |

### 15. Reproducibility artifacts saved today

| **Artifact** | **Purpose** |
| --- | --- |
| GSE224705_D5A_workspace.RData | Saved R workspace |
| GSE224705_D5A_analysis_history.Rhistory | Original R console history; 512 lines |
| GSE224705_D5A_analysis_history_readable.txt | Readable history copy |
| MMF_DEG_reproduced.rds | MMF DEG result |
| AZA_DEG_reproduced.rds | AZA DEG result |
| HC_DEG_reproduced.rds | HC DEG result |
| SOC_DEG_reproduced.rds | SOC DEG result |
| GSE224705_expression_reconstructed.rds | Expression matrix |
| GSE224705_metadata_reconstructed.rds | Metadata |

### 16. Suggested next roadmap

| **Step** | **Action** | **Why** |
| --- | --- | --- |
| 1 | Freeze current files | Preserve today's exact state |
| 2 | Reconstruct paper cohort exactly | Make DEG comparison truly like-for-like |
| 3 | Reconcile DEG discrepancy | Check cohort, preprocessing, contrast and adjustment |
| 4 | Align ML | Top-10 DEGs + patient-aware nested CV |
| 5 | Calculate AUROC/AUPRC | Standardized predictive benchmark |
| 6 | Add confidence intervals | Quantify uncertainty |
| 7 | Recover GSE135779 | Enable single-cell validation |
| 8 | Run CellChat/Hipathia | Mechanistic extension |
| 9 | Integrate findings | Bulk → cell → communication → target hypothesis |
| 10 | Freeze preprint | Methods, figures, code, limitations |

### 17. Effort and work-product assessment

Today's effort was substantially broader than a single DEG run. It included data reconstruction, metadata verification, author-code inspection, statistical reproduction, cohort debugging, clinical missing-data investigation, ML object inspection, GSEA interpretation, and preservation of the full interactive audit trail. The R history alone contains more than 1500 recorded console lines. This is evidence of substantial exploratory and reproducibility work, although the final publication code should be refactored into clean scripts rather than treating console history as production code.

### 18. Summary

We completed a reproducible audit/reconstruction of GSE224705 using the original paper and the authors' publicly available LNtherapy/pathMED code. Rather than replacing their methodology, we inspected and reused their core limma DEG framework and covariate structure, reconstructed the 319-sample expression/metadata dataset, rebuilt MMF/AZA/HC/SOC cohorts, and documented where the reconstructed cohorts differ from the final longitudinal cohorts reported in the paper. Our current DEG outputs are 222/4455/6/5 for MMF/AZA/HC/SOC versus the paper's 46/157/24/11. We are treating this as a reproducibility/cohort-processing question, not as an error claim. We also inspected the paper's nested patient-aware ML framework and existing RF/NB/KNN results; the current MMF RF model has MCC 0.626, balanced accuracy 0.825 and accuracy 0.897. Before making a direct performance claim, we will align the cohort/features/splits and calculate AUROC/AUPRC from valid out-of-fold probabilities. GSEA also shows significant treatment-specific cross-signature enrichment. The next stage is exact cohort reconciliation, the standardized ML benchmark, recovery of GSE135779 single-cell objects, and the downstream CellChat/Hipathia mechanistic analysis.

### 19. Primary references

https://pubmed.ncbi.nlm.nih.gov/38260685/

https://pmc.ncbi.nlm.nih.gov/articles/PMC10802741/

https://doi.org/10.21203/rs.3.rs-3783877/v1

https://www.ncbi.nlm.nih.gov/geo/query/acc.cgi?acc=GSE224705

https://www.ncbi.nlm.nih.gov/bioproject/PRJNA932236

https://github.com/dtordom/LNtherapy

https://github.com/jordimartorell/pathMED

https://bioc.r-universe.dev/pathMED/
